# Assessing the effect of a massive open online course (MOOC) on school water, sanitation, and hygiene improvements in the Philippines

**DOI:** 10.1101/2024.08.04.24311467

**Authors:** Kh. Shafiur Rahaman, Marvin Marquez, Sarah Bick, Dexter Galban, Habib Benzian, Bella Monse, Robert Dreibelbis

**Affiliations:** Department of Disease Control, London School of Hygiene and Tropical Medicine, London, UK; Fit for School Programme, Deutsche Gesellshaft fuer Internationale Zusammenarbeit (GIZ) GMBH, Makati City, Philippines; Department of Education, DepEd Complex, Meralco Avenue, Pasig City, Philippines; WHO Collaborating Center, Department Epidemiology & Health Promotion, College of Dentistry, New York University, New York, United States

## Abstract

Improving the water, sanitation, and hygiene (WASH) services in low-resources setting is a challenge. The Department of Education (DepEd) of the Philippines, supported by GIZ and UNICEF, runs the national WASH in schools program which promotes a stepwise approach to reach national WinS Standards and foster the institutionalization of WASH in the education sector. This includes national-level annual monitoring on WASH service levels in schools, information which is used to set target and allocation resources. Since 2019, the programme has also included a Massive Open Online Course (MOOC) for school staff. This platform provides uniform implementation guidance of WinS in the schools across the country. In this analysis, we use annual WASH monitoring data from the 2017/2018 (baseline) and 2021/2022 (endline) and compare this against school-level information on MOOC enrolment and completion. For each school in our analysis, we calculated baseline and endline overall and domain specific star ranking, a standardized 3-point composite measure of school WASH services adopted by DepEd. Linear regression models assessed the relationship between school staff participation in the MOOC and average change in star ranking between baseline and endline and logistic regression models were used to calculate the odds of improvement in star ranking between baseline and endline. Baseline and endline data were available for 28,779 schools. Of those, 5,980 schools had at least 1 teacher enrolled in the MOOC. Overall, MOOC participation was associated with improvements in both overall and domain specific star ranking, with larger improvements seen for hygiene services. The MOOC is a promising key component of the national WASH stategy complementing the annual monitoring process and warrants further investigation in the school management sector.

## Introduction

Adequate provision of water, sanitation, and hygiene (WASH) infrastructure and services together with improvements in WASH behaviours among school-going children are associated with a range of health and educational benefits [1]. Inadequate school WASH is associated with a range of infectious diseases that affect children’s overall health and can lead to reduced school attendance, reduced educational attainment, and a lower quality of life[2, 3]. On the contrary, evidence suggests that enhanced WASH services in schools can decrease student absence, reduce disease, and improve student well-being [1, 4-6].

Many schools, particularly in low resource settings, lack accessible and usable WASH facilities. Coverage of basic water, sanitation, and hygiene services remains low in lower-middle income countries like the Phillipines, with recent global estimates suggesting 1 in every 4 schools lacks access to basic water and basic sanitation services and almost half lack acess to basic hygiene services [7]. Sustained delivery, operation, and maintenance of WASH services in schools is also a challenge. Schools often lack the capacity to clean and maintain facilities, have inadequate budgets and resources. Irregular monitoring, lack of effective information-sharing systems, and weak accountability mechanisms contribute to limited service provision and poor sustainability [8-11].

In order to improve and sustain school WASH services, comprehensive approaches are recommended that include proper information flow, defined accountability mechanisms, tools to support school staff in the management and oversight of local WASH services, and clearly defined roles and repsonsiblities related to regular monitoring and system improvement [12]. This comprehensive approach to school WASH service provision has been central to the Fit for School Programme, a partnership between the Deutsche Gesellschaft für Internationale Zusammenarbeit (GIZ), the Southeast Asian Ministers of Education Organization (SEAMEO) and the Department of Education (DepEd) in the Philippines [13, 14]. Fit for School has supported the Philippine government on the design and implementation of a national school WASH programme. This programme, based on the GIZ/UNICEF “Three Star Approach for WASH in Schools” [15], promotes a stepwise approach to reach national WASH in schools standards and foster the institutionalisation of WASH in the education sector [16]. The programme uses a benchmarking system to categorize schools into specific star-rankings according to their level of service provision. The national programme aims to provide a clear roadmap by outlining improvement targets specific to each star level, establishing incentives for achieving those targets, and recognizing performance. The national indicators for the Philippines are aligned with both SDG targets (i.e., access to drinking water, gender-segregated usable toilets, and access to handwashing facilities with soap and water) [cite JMP], along with the nationally defined targets and standards. The program is monitored through through annual WASH surveys integrated in DepEd’s Education Information Management System (EMIS) and completed by all schools, with annual national data available from the 2017/2018 school year.

To support capacity for technical management and planning of WASH services at the school level and provide uniform implementation guideance; DepEd, the GIZ Regional Fit for School program and SEAMEO Innotech developed and launched a Massive Open Online Course (MOOC) [17] targeting the education sector’s teaching and managing workforce. The MOOC focuses on management and on-going maintenance of school WASH services and is aligned with the DepEd national WASH in schools policy. This digital learning platform is self-paced and caters to thousands of learners in batches. Integration of different activities (e.g., Facebook challenges, discussion forums, peer review etc.) encourages interaction among participants to share best practices and to motivate each other. Videos available throughout the course allow participants to use materials again with colleagues in their respective schools and divisions. DepEd maintains a database of all school staff who have enrolled in and completed the MOOC; participation in the MOOC counts towards the continuing education requirements of school staff. MOOCs are cited as a potential tool for providing education to a large number of learners, particularly learners who may not have time or resources to access other, more traditional forms of education. In low- and middle-income countries, MOOCs are a possible approach to filling critical human resource gaps necessary for continued professional education and capacity building, particularly among health professionals [18].

We explore the relationship between MOOC participation and changes in school-level WASH services between 2017 and 2021 by using two datasources for secondary data analysis – the national school-level WASH assesssments and the DepEd MOOC participation information..

## Methods

### Data sources

Two primary data sources were used in this analysis: annual school WASH assessments and DepED records on MOOC participation and completion. School WASH information is collected by three key stakeholder per year per school – the school head, a parent representative, and a community representative; the information is then uploaded to the EMIS portal. Following the Three Star approach, schools are assigned a star ranking according to compliance with nationally-defined indicators reflecting 17 elements of school WASH services. Individual indicators are converted into a composite measure that is then categorized into a 0, 1, 2, or 3-star rankings. A rank of 3-stars requires full compliance with national WASH in Schools standards (see Supplemental Information for DepEd’s WASH in Schools (WinS) Three-Star Approach Matrix, Table 1). In addition to the overall star ranking, DepEd provides domain specific star ranking for water, sanitation, and hygiene services. A detailed explanation of the star categories can be found elsewhere [16]. For the current analysis, we used national monitoring data from the 2017/2018 school year as baseline and compared this data from the 2021/2022 school year (endline). We replicated the national star ranking analysis, assigning schools an overall baseline and endline star ranking as well as domain specific star rankings for school water, sanitation, and hygiene services. MOOC data was taken from the DepED database of all school staff who enroll in and complete the school WASH MOOC which is offered twice annually. For a total of 4 “batches” or cohorts of learners it was possible to match MOOC participation data with the school WASH dataset based on school-name and school ID.

**Table 1:** School characteristics at baseline and MOOC participation between 2019 and 2021 (n=28,779)

|  |  |  |
| --- | --- | --- |
|  | N | % |
| **School Type** |  |  |
| Elementary | 24,130 | 84.1 |
| Secondary | 4,574 | 15.9 |
| **School size** |  |  |
| Small (0 – 440) | 20,875 | 72.7 |
| Medium (441 – 840) | 4,004 | 13.9 |
| Large (841- 1240) | 1,384 | 4.8 |
| Very large ($\geq 1241$) | 2,441 | 8.5 |
| **School location** |  |  |
| Urban | 1,363 | 4.9 |
| Semi-urban | 24,169 | 86.9 |
| Rural | 2,263 | 8.1 |
| **MOOC participation** |  |  |
| No school staff enrolled or completed | 22,799 | 79.2 |
| At least 1 school staff enrolled, no school staff completed | 1,264 | 4.4 |
| At least 1 school staff successfully completed | 4,716 | 16.4 |

### Data analysis

School overall and domain-specific star rankings were used as the primary outcomes in our analysis and assessed in multiple ways. First, we calculated changes in star ranking between baseline and endline for each school, resulting in an ordinal variable ranging from - 3 to 3. Second, we defined a binary variable reflecting any positive changes to the school’s overall or domain specific star ranking between baseline and endline (0 = no improvement or regression, 1 = improved star ranking).

Based on DepEd MOOC enrolment information, we defined three-level indicator of MOOC participation for each school. Schools were assigned 0 if no school staff had enrolled or completed the MOOC; a value of 1 if a school had at least 1 school staff enrolled in but did not complete the MOOC; and a value of 2 if the school had at least 1 staff member who completed the MOOC. MOOC participation was analysed as dummy variables in all subsequent analyses. For the purposes of our analysis, schools with no staff who enrolled in or compelted the MOOC were used as the reference group for our analysis.

The relationship between MOOC participation and changes in school WASH star ranking between 2017/2018 and 2021/2022 were assessed through multi-level regression models that included region as a random effect to account for geographic clustering. Multivariable linear regression models were used to compare MOOC enrolment and participation against changes in school and domain-specific star ranking between 2017/2018 and 2021/2022. For our binary indicator of improvement in star ranking, multivariable logistic regression models were used, and regression outputs were converted to odds ratios. To further explore the effect of the MOOC on the school’s with the highest need at baseline, this binary indicator of improvement in star ranking was restricted to those schools with a 0-star ranking at baseline.

All models were adjusted for covariates defined *a priori* with project partners, including type of schools (primary, secondary), school size, and location of the schools (urban/rural). Additional information on the economic status of the region was available for approximately 2/3 of all schools and fully adjusted model that includes the school’s economic region has been included in the supplemental information. Interaction tests were used to examine the differential effect of MOOC enrolment and participation by school level, location (urban/peri-urban/rural) and size.

Ethical approval for this secondary analysis was provided by the London School of Hygiene and Tropical Medicine ethics committee (Ethics ref no. 28381).

## Results

In the school year 2017/2018, 30,574 schools out of 46,654 invited submitted their WASH monitoring data. In the school year 2021/2022, 45,390 out of 48,533 schools submitted their WASH monitoring data on the digital platform. In total, complete baseline and endline school WASH data were available for 28,779 schools

### Characteristics of schools

Among schools with complete WinS monitoring data the majority were elementary schools (n=24,130; 84.1%), defined as small schools with less than 440 students (n=20,875, 72.7%) and semi-urban (n = 24,169, 87%). (Table 1).

Between 2019 and 2021, four batches of MOOC training were delivered; 15,741 school staff completed the course enrolment form with 15,558 school staff providing sufficient information to link their enrolment data with a specific school. After merging, there were 5,980 schools with at least 1 school staff who participated in the MOOC between 2019 and 2021 (20.8%). Among them, 4,716 schools (16.4%) had at least 1 school staff who completed the course and received the certificate. (Table 1). MOOC enrolment and completion status differed significantly by school level, school location and school size (See SI Table 2), with secondary schools, urban schools, and larger schools more likely to have staff who were enrolled in the MOOC or completed the MOOC (See SI Table 2). Adjustments for school type, location and size were made in all regression models.

**Table 2.** School domain-specific star ranking baseline and endline (N=28,779)

|  |  |  |  |  |
| --- | --- | --- | --- | --- |
|  | Baseline (2017/2018) |  | Endline (2021/2022) |  |
|  | n | % | n | % |
| **Water Star Ranking** |  |  |  |  |
| 0-star | 4,264 | 14.8 | 2,147 | 7.5 |
| 1-star | 10,724 | 37.4 | 7,100 | 24.7 |
| 2-star | 11,635 | 40.4 | 13,969 | 48.5 |
| 3-star | 2,156 | 7.5 | 5,563 | 19.3 |
| **Sanitation Star Ranking** |  |  |  |  |
| 0-star | 15,773 | 54.8 | 8,010 | 27.8 |
| 1-star | 8,715 | 30.3 | 7,418 | 25.8 |
| 2-star | 4,177 | 14.5 | 11,265 | 39.2 |
| 3-star | 114 | 0.4 | 2,086 | 7.3 |
| **Hygiene Star Ranking** |  |  |  |  |
| 0-star | 25,873 | 89.9 | 14,848 | 51.6 |
| 1-star | 1,789 | 6.2 | 3,129 | 10.9 |
| 2-star | 1,056 | 3.7 | 8,352 | 29.0 |
| 3-star | 61 | 0.2 | 2,450 | 8.5 |

### School Star Ranking

At baseline, 91% of schools had a 0-star ranking (n=26,173; 90.9%) for the composite WASH score, 3% had a 1-star ranking, 6% a 2-star ranking, and < 1% a 3-star ranking. At endline, schools with 0-star ranking reduced to 54.6% (n=15,717), 32% (n=9,169; 31.9%) of schools received 2-star ranking and approximately 9% of schools received a 3-star ranking (n=2,420). Only 2.5% of schools (n = 732) had a lower star ranking at endline compared to baseline, driven primarily by schools with 1- or 2-star rankings at baseline with zero star rankings at endline (n = 651). Details on the distribution of star ranking at baseline and endline are presented in Figure 1, with details on the specific specific star ranking at baseline and endline in Table 2.

**Figure 1:**
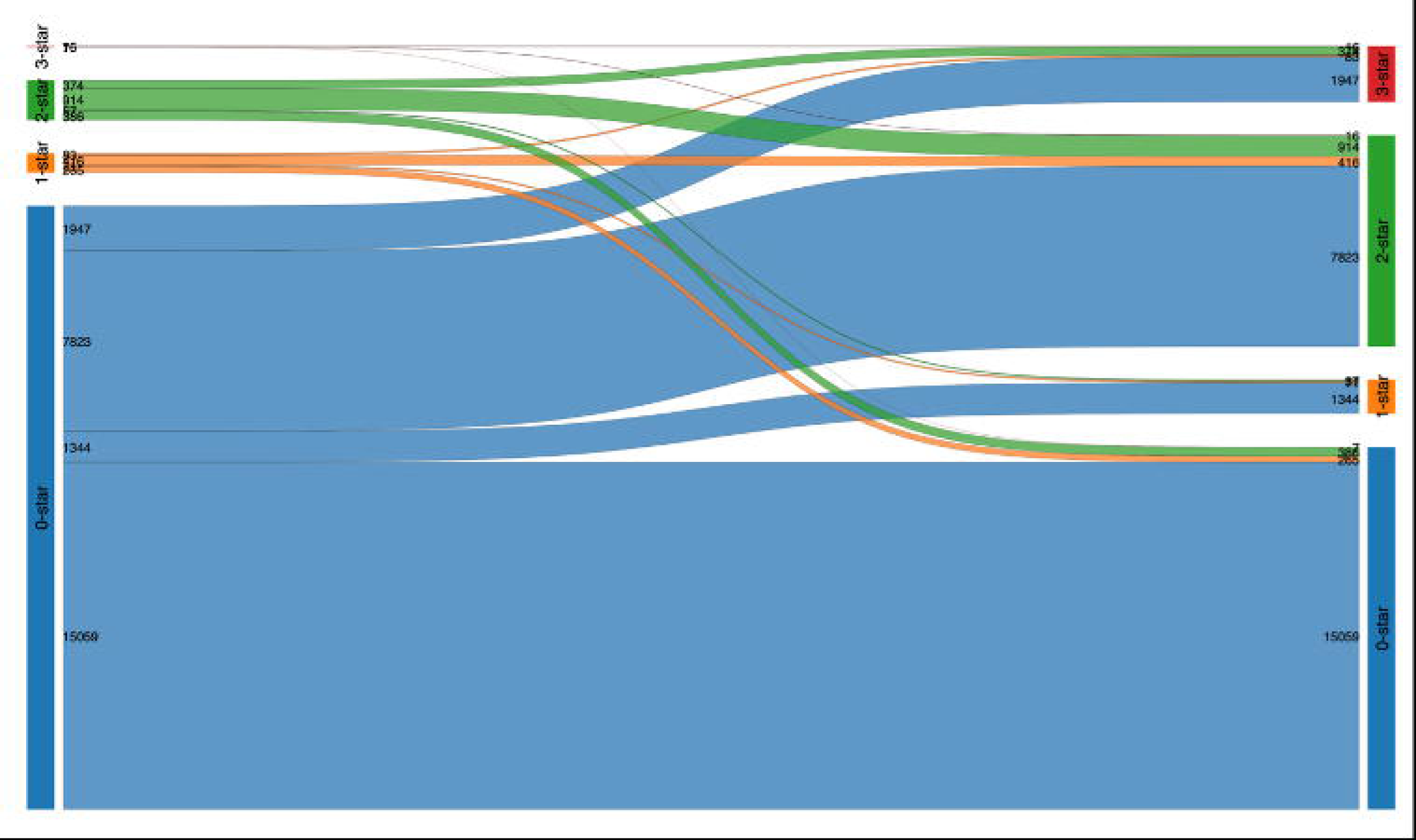
Sankey diagram of school WASH service level star ranking, 2017 / 2018 academic year to 2021 / 2022 academic year in the Philippines (n = 28,779)

### Regression results

In our adjusted regression models, schools with at least 1 school staff *enrolled* in the MOOC (referred at MOOC enrolment hereforeward) had a mean overall star ranking 0.14 points higher than schools that did not (β = 0.14; 95% confidence interval (CI): 0.08 – 0.20). Among schools with at least 1 school staff who *completed* the MOOC (referred to as MOOC completion), the mean overall star ranking between baseline and endline increased by 0.17 points (95% CI: 0.13 – 0.21) compared to schools with no school staff participation in the MOOC (Table 3). There was evidence of effect modification by school level (p=0.02) and school size (p=0.003),with with greater effects of MOOC completion among high schools and among schools with less than 440 pupils (See SI Table 3.1 and Table 3.3).

**Table 3:** Multivariable linear regression of changes in school star ranking between baseline and endline (n = 28,779)

|  |  |
| --- | --- |
| | $\beta$ (95% CI) |
| **Overall star ranking** |  |
| At least 1 school staff enrolled, no school staff completed | 0.14 (0.08 – 0.20) |
| At least 1 school staff completed the MOOC | 0.17 (0.13 – 0.21) |
| **Water Star Ranking** |  |
| At least 1 school staff enrolled, no school staff completed | 0.09 (0.04 – 0.15) |
| At least 1 school staff completed the MOOC | 0.08 (0.04 – 0.12) |
| **Sanitation Star Ranking** |  |
| At least 1 school staff enrolled, no school staff completed | 0.05 (-0.00 – 0.11) |
| At least 1 school staff completed the MOOC | 0.07 (0.03 – 0.10) |
| **Hygiene Star Ranking** |  |
| At least 1 school staff enrolled, no school staff completed | 0.13 (0.07 – 0.17) |
| At least 1 school staff completed the MOOC | 0.14 (0.09 – 0.18) |
| *all models adjusted for school level, location of the school, school size, and region. |  |

For domain-specific star rankings, MOOC enrolment was associated with a small increase in mean water star ranking between baseline and endline (β=0.09 (95% CI: 0.04 - 0.15)) after adjusting for covariates, with similar magnitude of change associated with MOOC completion. MOOC enrolment was not associated with changes in mean sanitation star ranking; however, MOOC completion was associated with a small improvement in sanitation star ranking (β=0.07 (95% CI: 0.03 - 0.10)) Changes in hygiene star ranking were associated with MOOC enrollment (β=0.13 (95% CI: 0.07 - 0.17)) and completion (β=0.14 (95% CI: 0.09 - 0.18)). (Table 3).

Logistic regression models of *any* improvement in star ranking between baseline and endline are showin in Table 4. The odds that a school would improve its overall star ranking between baseline and endline were 1.32 times higher in schools with at least 1 school staff enrolled in the MOOC compared to schools with no school staff enrolled (adjusted Odds Ratio (aOR): 1.32, 95% CI: 1.17 – 1.50)) and 1.57 times higher among schools were at least 1 staff member completed the MOOC (aOR: 1.57, 95% CI: 1.44 – 1.71) (Table 4). There was evidence of interaction between MOOC status and school size (p=0.029); the effect of MOOC enrolment on improvement in star ranking was greatest among small schools (<440 pupils; See SI Table 4.3).

**Table 4:** Adjusted ddds ratio of school improvement in overall and domain-specific star ranking between baseline and endline for MOOC enrolment and MOOC completion compared to no MOOC participation, all schools (n=28,779)*

| **Table 4: Adjusted odds ratio of school improvement in overall and domain-specific star ranking between baseline and endline for MOOC enrolment and MOOC completion compared to no MOOC participation, all schools (n=28,779)\*** |  |
| --- | --- |
|  | **Adjusted OR (95% CI)** |
| **Overall star ranking** |  |
| At least 1 school staff enrolled, but no school staff completed | 1.32 (1.17 – 1.50) |
| At least 1 school staff completed the MOOC | 1.57 (1.44 – 1.71) |
| **Water Star Ranking** |  |
| At least 1 school staff enrolled, but no school staff completed | 1.09 (0.98 – 1.23) |
| At least 1 school staff completed the MOOC | 1.13 (1.04 – 1.22) |
| **Sanitation Star Ranking** |  |
| At least 1 school staff enrolled, but no school staff completed | 1.09 (0.98 – 1.24) |
| At least 1 school staff completed the MOOC | 1.16 (1.07 – 1.25) |
| **Hygiene Star Ranking** |  |
| At least 1 school staff enrolled, but no school staff completed | 1.37 (1.21 – 1.55) |
| At least 1 school staff completed the MOOC | 1.52 (1.39 – 1.66) |
| *all models adjusted for school level, location of the school, school size, and region. |  |

For domain specific star-ranking, MOOC enrolment was not associated with a change in the odds of improvement in star ranking for water or sanitation. Schools with at least 1 staff who completed the MOOC had 13% higher odds of improving their water star ranking, 16% higher odds of improving their sanitation star ranking, and 52% higher odds of improving their hygiene star ranking.

*Sub-analysis*:

Among schools with 0-stars at baseline (n = 26,173) schools with at least 1 school staff enrolled in the MOOC had 1.39 times higher odds of improving star ranking between baseline and endline (aOR 1.39, 95% CI: 1.22– 1.59). The odds of a school improving it’s overall star ranking from 0 between baseline and endline was 1.92 times higher among schools with at least 1 staff member completing the MOOC compared to schools than schools with no school staff participating in the WinS MOOC (aOR 1.92, 95 CI: 1.75 – 2.12) (Table 5). There was evidence of interaction between MOOC status and school size (p=0.029); the effect of MOOC enrolment on improvement from zero stars was greatest among small schools (See SI Table 5.3).

**Table 5:** Adjusted ddds ratio of school improvement in overall and domain-specific star ranking between baseline and endline for MOOC enrolment and MOOC completion compared to no MOOC participation amond schools with 0-stars at baseline*.

|  | **Adjusted OR (95% CI)** |
| --- | --- |
| **Overall star ranking (n = 26,173)** |  |
| At least 1 school staff enrolled, no school staff completed | 1.39 (1.22 – 1.59) |
| At least 1 school staff completed the MOOC | 1.92 (1.75 – 2.12) |
| **Water Star Ranking (n = 4,264)** |  |
| At least 1 school staff enrolled, no school staff completed | 1.35 (0.79 – 2.29) |
| At least 1 school staff completed the MOOC | 1.49 (1.05 – 2.13) |
| **Sanitation Star Ranking (n= 15, 773)** |  |
| At least 1 school staff enrolled, no school staff completed | 1.28 (1.03 – 1.58) |
| At least 1 school staff completed the MOOC | 1.56 (1.35 – 1.79) |
| **Hygiene Star Ranking (n= 25,873)** |  |
| At least 1 school staff enrolled, no school staff completed | 1.42 (1.24 – 1.62) |
| At least 1 school staff completed the MOOC | 1.76 (1.59 – 1.93) |
| *all models adjusted for school level, location of the school, school size, and region. |  |

To assess the robustness of our analysis, we re-ran all models with information on the economic status of school locations, resulting in models with only 20,799 records. In this additional analysis (SI Table 6), the significance and magnitude of change remained similar across most outcomes of interest with minor exceptions to the domain specific star rankings. Specifically, mean changes in sanitation star ranking were no longer associated with MOOC completion, and changes in the odds of improvement between baseline and endline in water and sanitation star ranking were no longer associated with MOOC enrolment and MOOC completion. However, results from this analysis should be interpreted with caution due to the large loss of data.

## Discussion

We note a general improvement in school WASH services over the period of our analysis, a period in which DepEd implemented a large-scale WASH in schools programme. Over 40% of schools with a 0-star ranking in 2017 had improved their overall WASH services, many to 2-stars or higher. Specific to this analsyis, we assessed the relationship between participation in a MOOC as a capacity-strengthening digital learning platform and school-level changes in WASH service in a national sample of schools over a five-year period. We found improvements in overall school WASH service provision and improvements in the specific domains of school WASH associated with both participation and completion in the DepEd-managed MOOC. Our findings suggest that online digital learning platforms, as part of a comprehensive school WASH programme, can have positive effects at a national scale.

The association between MOOC participation and completion and improvements in star rankings were greater among schools with higher needs ( ie. having no access to basic WASH services) at baseline. However, across all analyses, improvements were modest; schools with school staff enrolled improved by 0.14 points more than their counterparts on a 0–3 scale and few schools were fully aligned with national WASH in Schools standard (3-star ranking) at endline. The MOOC therefore might play a critical role in progressing schools up the JMP WASH service ladders, particularly from a lower starting point, or sustaining improvements at the ‘basic’ level. This aspect is of particular relevance as more than 90% of schools started at an overall 0-star level.

We observed the largest changes in handwashing service delivery at schools. Improvements in school water and sanitation – even to meet basic standards – likely require significant investments in financial and human resources which take longer time to be integrated into government planning, budget allocation and construction procedures. Moreover, the MOOC intervention was targeted to strengthen the existing WASH management capacities of the education system, while investment into infrastructure was explicitly not within the scope of the intervention. Our sample had relatively robust water access at baseline (only 15% of schools in our sample had a 0-star water ranking at baseline); and the necessary water supply improvements required to enable effective hand hygiene were in place in the majority of our schools. Improvement in hygiene services may require management capacities and small resource investments, particularly when low-cost handwashing infrastructure such as the WASHALOT [19] system promoted in the Philippines is installed. Future interventions may consider the development of a specific MOOC targeting decision makers at national/ subnational levels in charge of financial planning and resource allocation in order to increase investment in WASH infrastructure.

Sunstained provision of WASH services in schools is a necessary component of any school WASH programme and essential for WASH services to achieve their intended impacts on health and education [1, 20]. However, findings from experimental studies of service provision models are mixed [21-24]. Based on a review of published barriers and enablers and service delivery intervention, Pu and colleagues propose three necessary conditions for sustainable delivery of school WASH services: resources (human, financial, physical, etc), timely and credibly information, and accountability mechanisms [12]. The comprehensive school WASH strategy, including the MOOC for school staff, implemented by DepEd in the Philippines with technical support from development partners, serves as a potential model for addressing these three components at a national level. The policy and program connect the accountability mechanism of annual reporting with financial rewards and incentives. They also enhance the human and technical capacity of schools through innovative training models, incentivizing school staff to complete these programs.

Published analyses of MOOCs specific to the WASH sector are limited [25, 26], with available studies focusing on reach, learner retention, and learner satisfaction. To our knowledge, this is the first analysis that links MOOC participation with changes in WASH service provision. MOOCs often suffer from low rates of completion [25-28]. Linking MOOC completion with continuing education credits may have contributed to higher completion rates in the DepEd programme than commonly observed in other studies. We found improvements in school WASH services associated with both enrolment and completion of the MOOC programme, suggesting that even limited engagement with the MOOC may have been sufficient to improve institution-wide engagement and produce change to school WASH services. Alternatively, staff who were more motivated to engage in school WASH activities may have also been more likely to participate in the MOOC.

The strengths of the study include its national focus, the large sample size, and the ability to compare WASH status before and after the intervention in the same schools over a five-year period. However, we relied on crude measures of MOOC participation and only two data points reflecting school WASH conditions over the observation period. The study uses self-reported data on school WASH services available in DepEd’s EMIS. The EMIS data collection system aims to reduce bis through joint data gathering by 3 members of the school community; the reported WASH data are validated at multiple levels of the DepEd administration as well as through technical support by GIZ and the WHO/UNICEF Joint Monitoring Programme. It is unclear whether a MOOC, successful in the context of the Philippines education sector, would have effects generalisable to other regions and political contexts; particularly given the high rates of mobile phone coverage and internet use in the Phillippines. Further randomised trials that include independent observations of facilities and longitudinal follow-up with more frequent measurements could help to assess the trajectory of change in WASH services in schools and attribute effects to such capacity building interventions.

## Conclusion

This national-scale study found that a Massive Open Online Course as an educational intervention for school staff was able to support the sustained delivery and improvement of WASH services in schools in the Philippines over a five-year period. At the school level, having any school staff enrolled in the programme was associated with improvements in school WASH status in general; further research is needed to confirm causal links between these improvements and MOOC participation. Findings indicate that a MOOC offers a realistic approach for large-scale capacity development at low-cost and is able to support and complement national WASH in Schools programmes.

## Supporting information

Supplmental Tables MOOC

## Data Availability

Data in the present work are the property of Department of Education (DepEd), Government of Philippines. Please contact Assistant Secretary Dexter Galban for access to the data.

