## Supplementary material for "Assessing the effect of a massive open online course (MOOC) on school water, sanitation, and hygiene improvements in the Philippines": Supplmental Tables MOOC

Supporting Information

| **SI: Table 1 DepEd’s WASH in Schools Three-Star Approach** |
| --- |
| **Star Rating** |
| **3-star**   - All school water, sanitation, and hygiene facilities and promotion meet or exceed national standards |
| **2-star**   - Hygiene education and facilities to promote handwashing with soap after toilet use available - Improved sanitation facilities available, along with facilities and education related to menstrual hygiene management - Low-cost point of use water treatment in schools routinely completed |
| **1-star**   - Daily group handwashing with soap completed, normally before the school mean - Daily supervised cleaning of toilets and provision of water and soap - At least one usable toilet for girls and one for boys - No open defecation |
| **0-star**   - No or limited hygiene promotion - Does not meet all five of the following: i) no safe drinking water available, ii) no gender segregated toilets, iii)no group handwashing facilities with both water and soap, iv) no daily group handwashing activities conducted, and v) no access to emergency pads |

| **SI: Table 2 Chi-squared test between MOOC Participation and School Type (N = 28, 779)** | | | | | |
| --- | --- | --- | --- | --- | --- |
| **School characteristics** | **MOOC Participation, n (%)** | | | **Chi^2^** | **P value** |
|  | No participation | Participated, not completed | Participated and completed |  |  |
| **School type** |  |  |  | 503. 8 | **<0.001** |
| Elementary | 19642 (81.4) | 855 (3.5) | 3633 (15.1) |  |  |
| Secondary | 3098 (67.7) | 407 (8.9) | 1069 (23.4) |  |  |
| **School location** |  |  |  | 737.0 | **<0.001** |
| Rural | 1126 (82.6) | 42 (3.1) | 195 (14.3) |  |  |
| Peri-urban | 19450 (80.5) | 965 (4.0) | 3754 (15.5) |  |  |
| Urban | 1280 (56.6) | 244 (10.8) | 739 (32.7) |  |  |
| **School size** |  |  |  | 1300 | **<0.001** |
| Small (<440 pupils) | 17470 (83.7) | 623 (3.0) | 2782 (13.3) |  |  |
| Medium (441–840 pupils) | 2960 (73.9) | 229 (5.7) | 815 (20.4) |  |  |
| Large (841–1240 pupils) | 919 (66.4) | 111 (8.0) | 354 (25.6) |  |  |
| Very large (>1241 pupils) | 1391 (57.0) | 299 (12.3) | 751 (30.8) |  |  |

| **SI Table 3 Multivariable linear regression of changes in school star ranking between baseline and endline (N = 28, 779)** | | | | | | | | |
| --- | --- | --- | --- | --- | --- | --- | --- | --- |
|  | **Overall School WASH Star Ranking** | | **Water** | | **Sanitation** | | **Hygiene** | |
|  | β | 95% CI  (Lower, Upper) | β | 95% CI  (Lower, Upper) | β | 95% CI  (Lower, Upper) | β | 95% CI  (Lower, Upper) |
| **MOOC Participation** | | | | | | | | |
| *No school participated* | Ref | | Ref | | Ref | | Ref | |
| *At least 1 teacher enrolled, but no teacher completed* | 0.14 | 0.08, 0.20 | 0.09 | 0.35, 0.15 | 0.05 | -0.00, 0.11 | 0.12 | 0.07, 0.17 |
| *At least 1 teacher completed the MOOC* | 0.17 | 0.13, 0.21 | 0.08 | 0.04, 0.12 | 0.07 | 0.03, 0.10 | 0.14 | 0.09, 0.18 |
| **School Type** |  |  |  |  |  |  |  |  |
| *Elementary* | Ref | | Ref | | Ref | | Ref | |
| *Secondary* | 0.03 | -0.00, 0.07 | 0.10 | 0.07, 0.14 | -0.08 | -0.02, 0.08 | -0.03 | -0.07, 0.00 |
| **Location of Schools** | | | | | | | | |
| *Urban* | Ref | | Ref | | Ref | | Ref | |
| *Sei-urban* | 0.11 | 0.05, 0.16 | 0.03 | 0.00, 0.11 | 0.03 | -0.02, 0.08 | 0.11 | 0.06, 0.17 |
| *Rural* | -0.03 | -0.19, 0.12 | 0.04 | -0.03, 0.11 | 0.02 | -0.05, 0.09 | 0.05 | -0.03, 0.13 |
| **School Size** | | | | | | | | |
| *Small* | Ref | | Ref | | Ref | | Ref | |
| *Medium* | 0.28 | 0.25, 0.32 | 0.03 | -0.00, 0.07 | 0.20 | 0.17, 0.24 | 0.21 | 0.17, 0.25 |
| *Large* | 0.28 | 0.22, 0.33 | 0.04 | -0.01, 0.09 | 0.19 | 0.14, 0.25 | 0.21 | 0.15, 0.27 |
| *Very Large* | 0.36 | 0.30, 0.41 | -0.01 | -0.06, 0.04 | 0.13 | 0.08, 0.17 | 0.23 | 0.18, 0.28 |
| **Region** | 0.10 | 0.05, 0.21 | 0.01 | 0.00, 0.04 | 0.05 | 0.02, 0.09 | 0.11 | 0.05, 0.21 |

| **SI Table 3.1 Stratum-specific odds ratios from Multivariable Linear Regression (changes in star ranking) with interaction between MOOC participation and school type. (N=27, 795)** | | | | |
| --- | --- | --- | --- | --- |
|  | | OR | 95% CI | P value |
| **Overall star ranking** | |  |  |  |
| *Elementary schools* | |  |  |  |
|  | At least 1 school staff enrolled, but no school staff completed | 0.17 | 0.10-0.25 | <0.001 |
|  | At least 1 school staff completed the MOOC | 0.15 | 0.11-0.19 | <0.001 |
| *High schools* | | | | |
|  | At least 1 school staff enrolled, but no school staff completed | 0.09 | -0.02 – 0.20 | 0.094 |
|  | At least 1 school staff completed the MOOC | 0.24 | 0.17 – 0.31 | <0.001 |
| Likelihood ratio test for interaction [MOOC participation/completion and school type] | |  |  | 0.020 |

| **SI Table 3.2** **Stratum-specific odds ratios from Multivariable Linear Regression (Changes in star ranking) with interaction between MOOC participation and school Location. (N=27, 795)** | | | | |
| --- | --- | --- | --- | --- |
|  |  | OR | 95% CI | P value |
| **Overall star ranking** | |  |  |  |
| *Rural* | |  |  |  |
|  | At least 1 school staff enrolled, but no school staff completed | 0.57 | 0.26-0.88 | <0.001 |
|  | At least 1 school staff completed the MOOC | 0.20 | 0.04-0.35 | 0.013 |
| *Peri-urban* | | | | |
|  | At least 1 school staff enrolled, but no school staff completed | 0.15 | 0.08-0.21 | <0.001 |
|  | At least 1 school staff completed the MOOC | 0.16 | 0.12-0.21 | <0.001 |
| *Urban* | | | | |
|  | At least 1 school staff enrolled, but no school staff completed | 0.06 | -0.08 - 0.19 | 0.428 |
|  | At least 1 school staff completed the MOOC | 0.18 | 0.09 – 0.28 | <0.001 |
| Likelihood ratio test for interaction [MOOC participation/completion and school location] | |  |  | 0.055 |

| **SI Table 3.3 Stratum-specific odds ratios from Multivariable Linear Regression (Changes in star ranking) with the interaction between MOOC participation and school Size. (N=27, 795)** | | | | |
| --- | --- | --- | --- | --- |
|  |  | OR | 95% CI | P value |
| *Small schools (<440 pupils)* | |  |  |  |
|  | At least 1 school staff enrolled, but no school staff completed | 0.23 | 0.15-0.31 | <0.001 |
|  | At least 1 school staff completed the MOOC | 0.13 | 0.08-0.18 | <0.001 |
| *Medium schools (441–840 pupils)* | | | | |
|  | At least 1 school staff enrolled, but no school staff completed | 0.05 | -0.08 – 0.19 | 0.076 |
|  | At least 1 school staff completed the MOOC | 0.25 | 0.16-0.33 | <0.001 |
| *Large schools (841–1240 pupils)* | | | | |
|  | At least 1 school staff enrolled, but no school staff completed | 0.18 | -0.03 – 0.38 | 0.088 |
|  | At least 1 school staff completed the MOOC | 0.26 | 0.13-0.38 | <0.001 |
| *Very large schools (>1241 pupils)* | | | | |
|  | At least 1 school staff enrolled, but no school staff completed | 0.03 | -0.10-0.15 | 0.689 |
|  | At least 1 school staff completed the MOOC | 0.17 | 0.08-0.26 | <0.001 |
| **Likelihood ratio test for interaction [MOOC participation/completion and school size]** | |  |  | 0.003 |

| **SI Table 4 Multivariable logistic regression of improvements in school star ranking between baseline and endline (n = 28, 779)** | | | | | | | | |
| --- | --- | --- | --- | --- | --- | --- | --- | --- |
|  | **Overall School WASH Star Ranking** | | **Water** | | **Sanitation** | | **Hygiene** | |
|  | OR | 95% CI  (Lower, Upper) | OR | 95% CI  (Lower, Upper) | 0R | 95% CI  (Lower, Upper) | OR | 95% CI  (Lower, Upper) |
| **MOOC Participation** | | | | | | | | |
| *No school participated* | Ref | | Ref | | Ref | | Ref | |
| *At least 1 teacher enrolled, but no teacher completed* | 1.32 | 1.17, 1.50 | 1.09 | 0.98, 1.23 | 1.09 | 0.98, 1.24 | 1.37 | 1.21, 1.55 |
| *At least 1 teacher completed the MOOC* | 1.57 | 1.44, 1.71 | 1.13 | 1.04, 1.22 | 1.16 | 1.07, 1.25 | 1.52 | 1.39, 1.66 |
| **Type of School** |  |  |  |  |  |  |  |  |
| *Elementary* | Ref | | Ref | | Ref | | Ref | |
| *Secondary* | 0.97 | 0.89, 1.05 | 1.22 | 1.14, 1.31 | 1.14 | 1.07, 1.24 | 0.78 | 0.72, 0.85 |
| **Location of Schools** | | | | | | | | |
| *Urban* | Ref | | Ref | | Ref | | Ref | |
| *Sei-urban* | 1.28 | 1.12, 1.46 | 1.11 | 0.99, 1.25 | 0.99 | 0.88, 1.11 | 1.22 | 1.17, 1.51 |
| *Rural* | 1.14 | 0.96, 1.35 | 1.06 | 0.91, 1.23 | 0.94 | 0.80, 1.09 | 1.22 | 1.04, 1.44 |
| **School Size** | | | | | | | | |
| *Small* | Ref | | Ref | | Ref | | Ref | |
| *Medium* | 1.72 | 1.59, 1.86 | 1.05 | 0.99, 1.13 | 1.54 | 1.43, 1.66 | 1.56 | 1.45, 1.69 |
| *Large* | 1.69 | 1.49, 1.92 | 1.07 | 0.95, 1.19 | 1.41 | 1.25, 1.59 | 1.59 | 1.40, 1.80 |
| *Very Large* | 2.05 | 1.83, 2.28 | 1.91 | 0.91, 1.11 | 1.21 | 1.09, 1.35 | 1.82 | 1.63, 2.03 |
| **Region** | 0.47 | 0.24, 0.95 | 0.06 | 0.03, 0.13 | 0.19 | 0.09, 0.39 | 0.46 | 0.23, 0.92 |

| **SI Table 4.1 Stratum-specific odds ratios from Multivariable Logistic Regression (Improvement in star ranking) with interaction between MOOC participation and school type. (N=27, 795)** | | | | |
| --- | --- | --- | --- | --- |
|  | | OR | 95% CI | P value |
| **Overall star ranking** | |  |  |  |
| *Elementary schools* | |  |  |  |
|  | At least 1 school staff enrolled, but no school staff completed | 1.39 | 1.19 – 1.61 | < 0.001 |
|  | At least 1 school staff completed the MOOC | 1.52 | 1.38 – 1.67 | < 0.001 |
| *High schools* | | | | |
|  | At least 1 school staff enrolled, but no school staff completed | 1.22 | 0.97-1.52 | 0.078 |
|  | At least 1 school staff completed the MOOC | 1.75 | 1.48-2.07 | <0.001 |
| **Likelihood ratio test for interaction [MOOC participation/completion and school type]** | |  |  | 0.146 |

| **SI Table 4.2** **Stratum-specific odds ratios from Multivariable Logistic Regression (Improvement in star ranking) with interaction between MOOC participation and school Location. (N=27, 795)** | | | | |
| --- | --- | --- | --- | --- |
|  |  | OR | 95% CI | P value |
| **Overall star ranking** | |  |  |  |
| *Rural* | |  |  |  |
|  | At least 1 school staff enrolled, but no school staff completed | 3.33 | 1.67-6.62 | 0.001 |
|  | At least 1 school staff completed the MOOC | 1.82 | 1.26-2.65 | 0.001 |
| *Peri-urban* | | | | |
|  | At least 1 school staff enrolled, but no school staff completed | 1.32 | 1.14-1.51 | <0.001 |
|  | At least 1 school staff completed the MOOC | 1.56 | 1.42-1.72 | <0.001 |
| *Urban* | | | | |
|  | At least 1 school staff enrolled, but no school staff completed | 1.14 | 0.85-1.53 | 0.366 |
|  | At least 1 school staff completed the MOOC | 1.49 | 1.21-1.83 | <0.001 |
| **Likelihood ratio test for interaction [MOOC participation/completion and school location]** | |  |  | 0.072 |

| **SI Table 4.3 Stratum-specific odds ratios from Multivariable Logistic Regression (Improvement in star ranking) with the interaction between MOOC participation and school Size. (N=27, 795)** | | | | |
| --- | --- | --- | --- | --- |
|  |  | OR | 95% CI | P value |
| *Small schools (<440 pupils)* | |  |  |  |
|  | At least 1 school staff enrolled, but no school staff completed | 1.61 | 1.36-1.93 | <0.001 |
|  | At least 1 school staff completed the MOOC | 1.51 | 1.36-1.68 | <0.001 |
| *Medium schools (441–840 pupils)* | | | | |
|  | At least 1 school staff enrolled, but no school staff completed | 0.99 | 0.75-1.32 | 0.993 |
|  | At least 1 school staff completed the MOOC | 1.72 | 1.43-2.07 | <0.001 |
| *Large schools (841–1240 pupils)* | | | | |
|  | At least 1 school staff enrolled, but no school staff completed | 1.38 | 0.90-2.10 | 0.130 |
|  | At least 1 school staff completed the MOOC | 1.71 | 1.29-2.28 | <0.001 |
| *Very large schools (>1241 pupils)* | | | | |
|  | At least 1 school staff enrolled, but no school staff completed | 1.07 | 0.82-1.39 | 0.619 |
|  | At least 1 school staff completed the MOOC | 1.50 | 1.22-1.84 | <0.001 |
| **Likelihood ratio test for interaction [MOOC participation/completion and school size]** | |  |  | 0.0291 |

| **SI Table 5 Multivariable logistic regression of improvements in school star ranking from zero between baseline and endline** | | | | | | | | |
| --- | --- | --- | --- | --- | --- | --- | --- | --- |
|  | **Overall School WASH Star Ranking**  (n=26, 173) | | **Water**  (n=4, 264) | | **Sanitation**  (n=15, 773) | | **Hygiene**  (n= 25, 873) | |
|  | OR | 95% CI  (Lower, Upper) | OR | 95% CI  (Lower, Upper) | 0R | 95% CI  (Lower, Upper) | OR | 95% CI  (Lower, Upper) |
| **MOOC Participation** | | | | | | | | |
| *No school participated* | Ref | | Ref | | Ref | | Ref | |
| *At least 1 teacher enrolled, but no teacher completed* | 1.39 | 1.22, 1.59 | 1.35 | 0.79, 2.29 | 1.28 | 1.03, 1.58 | 1.42 | 1.24, 1.62 |
| *At least 1 teacher completed the MOOC* | 1.92 | 1.75, 2.12 | 1.49 | 1.05, 2.13 | 1.56 | 1.35, 1.79 | 1.76 | 1.59, 1.93 |
| **Type of School** |  |  |  |  |  |  |  |  |
| *Elementary* | Ref | | Ref | | Ref | | Ref | |
| *Secondary* | 0.89 | 0.82, 0.97 | 1.18 | 0.87, 1.61 | 2.94 | 2.48, 3.48 | 0.73 | 0.67, 0.79 |
| **Location of Schools** | | | | | | | | |
| *Urban* | Ref | | Ref | | Ref | | Ref | |
| *Sei-urban* | 1.32 | 1.14, 1.50 | 1.42 | 1.03, 1.98 | 1.02 | 0.87, 1.21 | 1.39 | 1.21, 1.59 |
| *Rural* | 1.21 | 1.01, 1.44 | 2.14 | 0.99, 4.68 | 1.22 | 0.95, 1.56 | 1.28 | 1.07, 1.52 |
| **School Size** | | | | | | | | |
| *Small* | Ref | | Ref | | Ref | | Ref | |
| *Medium* | 1.85 | 1.70, 2.02 | 1.67 | 1.18, 2.38 | 2.02 | 1.79, 2.29 | 1.62 | 1.49, 1.77 |
| *Large* | 1.85 | 1.62, 2.12 | 1.34 | 0.71, 2.53 | 2.48 | 1.96, 3.14 | 1.63 | 1.43, 1.86 |
| *Very Large* | 2.41 | 2.14, 2.72 | 3.66 | 1.29, 10.36 | 3.23 | 2.50, 4.16 | 1.95 | 1.73, 2.19 |
| **Region** | 0.72 | 0.36, 1.43 | 0.29 | 0.10, 0.81 | 0.59 | 0.28, 1.23 | 0.61 | 0.31, 1.21 |

| **SI Table 5.1 Stratum-specific odds ratios from Multivariable Logistic Regression (Improvement in star ranking from zero) with interaction between MOOC participation and school type. (N=25,278)** | | | | |
| --- | --- | --- | --- | --- |
|  | | OR | 95% CI | P value |
| **Overall star ranking** | |  |  |  |
| *Elementary schools* | |  |  |  |
|  | At least 1 school staff enrolled, but no school staff completed | 1.51 | 1.28-1.78 | < 0.001 |
|  | At least 1 school staff completed the MOOC | 1.90 | 1.71-2.12 | < 0.001 |
| *High schools* | | | | |
|  | At least 1 school staff enrolled, but no school staff completed | 1.20 | 0.96-1.53 | 0.114 |
|  | At least 1 school staff completed the MOOC | 1.97 | 1.64-2.37 | <0.001 |
| **Likelihood ratio test for interaction [MOOC participation/completion and school type]** | |  |  | 0.275 |

| **SI Table 5.2 Stratum-specific odds ratios from Multivariable Logistic Regression (Improvement in star ranking) with interaction between MOOC participation and school Location. (N=25,278)** | | | | |
| --- | --- | --- | --- | --- |
|  |  | OR | 95% CI | P value |
| **Overall star ranking** | |  |  |  |
| *Rural* | |  |  |  |
|  | At least 1 school staff enrolled, but no school staff completed | 3.02 | 1.46-6.21 | 0.003 |
|  | At least 1 school staff completed the MOOC | 1.58 | 1.02-2.45 | 0.040 |
| *Peri-urban* | | | | |
|  | At least 1 school staff enrolled, but no school staff completed | 1.39 | 1.19-1.62 | <0.001 |
|  | At least 1 school staff completed the MOOC | 1.93 | 1.73-2.14 | <0.001 |
| *Urban* | | | | |
|  | At least 1 school staff enrolled, but no school staff completed | 1.24 | 0.90-1.70 | 0.192 |
|  | At least 1 school staff completed the MOOC | 1.98 | 1.57-2.50 | <0.001 |
| **Likelihood ratio test for interaction [MOOC participation/completion and school location]** | |  |  | 0.183 |

| **SI Table 5.3 Stratum-specific odds ratios from Multivariable Logistic Regression (Improvement in star ranking) with the interaction between MOOC participation and school Size. (N=25,278)** | | | | |
| --- | --- | --- | --- | --- |
|  |  | OR | 95% CI | P value |
| *Small schools (<440 pupils)* | |  |  |  |
|  | At least 1 school staff enrolled, but no school staff completed | 1.71 | 1.42-2.07 | <0.001 |
|  | At least 1 school staff completed the MOOC | 1.86 | 1.64-2.10 | <0.001 |
| *Medium schools (441–840 pupils)* | | | | |
|  | At least 1 school staff enrolled, but no school staff completed | 1.00 | 0.73-1.36 | 0.986 |
|  | At least 1 school staff completed the MOOC | 2.06 | 1.67-2.54 | <0.001 |
| *Large schools (841–1240 pupils)* | | | | |
|  | At least 1 school staff enrolled, but no school staff completed | 1.51 | 0.96-2.37 | 0.076 |
|  | At least 1 school staff completed the MOOC | 2.22 | 1.61-3.07 | <0.001 |
| *Very large schools (>1241 pupils)* | | | | |
|  | At least 1 school staff enrolled, but no school staff completed | 1.13 | 0.83-1.50 | 0.413 |
|  | At least 1 school staff completed the MOOC | 1.82 | 1.44-2.30 | <0.001 |
| **Likelihood ratio test for interaction [MOOC participation/completion and school size]** | |  |  | 0.039 |

| **SI Table 6 Multivariable linear regression of changes in school star ranking between baseline and endline (with economic status of school locations (n = 20, 779)** | | | | | | | | |
| --- | --- | --- | --- | --- | --- | --- | --- | --- |
|  | **Overall School WASH Star Ranking** | | **Water** | | **Sanitation** | | **Hygiene** | |
|  | β | 95% CI  (Lower, Upper) | β | 95% CI  (Lower, Upper) | β | 95% CI  (Lower, Upper) | β | 95% CI  (Lower, Upper) |
| **MOOC Participation** | | | | | | | | |
| *No school participated* | Ref | | Ref | | Ref | | Ref | |
| *At least 1 teacher enrolled, but no teacher completed* | **0.15** | 0.07, 9.22 | **0.12** | 0.04, 0.19 | **0.08** | 0.01, 0.15 | **0.15** | 0.08, 0.22 |
| *At least 1 teacher completed the MOOC* | **0.11** | 0.06, 0.16 | **0.06** | 0.01, 0.11 | 0.04 | -0.00, 0.09 | **0.07** | 0.02, 0.12 |
| **Type of School** |  |  |  |  |  |  |  |  |
| *Elementary* | Ref | | Ref | | Ref | | Ref | |
| *Secondary* | 0.04 | -0.01, 0.08 | **0.08** | 0.04, 0.13 | **-0.07** | -0.11, -0.03 | -0.02 | -0.06, 0.03 |
| **Location of Schools** | | | | | | | | |
| *Urban* | Ref | | Ref | | Ref | | Ref | |
| *Sei-urban* | **0.08** | 0.02, 0.14 | 0.06 | -0.00, 0.11 | 0.00 | -0.05, 0.06 | **0.09** | 0.03, 0.15 |
| *Rural* | -0.09 | -0.20, 0.03 | -0.06 | -0.17, 0.05 | -0.08 | -0.19, 0.03 | -0.09 | -0.21, 0.02 |
| **School Size** | | | | | | | | |
| *Small* | Ref | | Ref | | Ref | | Ref | |
| *Medium* | **0.28** | 0.24, 0.33 | 0.03 | -0.02, 0.07 | **0.21** | 0.16, 0.23 | **0.21** | 0.17, 0.25 |
| *Large* | **0.29** | 0.21, 0.36 | 0.04 | -0.03, 0.12 | **0.19** | 0.12, 0.26 | **0.22** | 0.14, 0.29 |
| *Very Large* | **0.36** | 0.29, 0.42 | 0.02 | -0.04, 0.09 | **0.19** | 0.13, 0.26 | **0.23** | 0.16, 0.29 |
| **Economic status of school locations** | | | | | | | | |
| *Income class 1 (High)* | Ref | | Ref | | Ref | | Ref | |
| *Income class 2* | 0.03 | -0.02, 0.07 | 0.04 | -0.01, 0.08 | 0.03 | -0.02, 0.07 | 0.02 | -0.03, 0.06 |
| *Income class 3* | -0.03 | -0.07, 0.01 | -0.03 | -0.08, 0.00 | -0.03 | -0.06, 0.01 | -0.03 | -0.07, 0.01 |
| *Income class 4* | **0.05** | 0.01, 0.09 | 0.06 | 0.02, 0.10 | 0.02 | -0.01, 0.06 | 0.04 | 0.00, 0.08 |
| *Income class 5* | -0.01 | -0.06, 0.04 | **0.07** | 0.02, 0.13 | -0.03 | -0.08, 0.02 | -0.01 | -0.06, 0.04 |
| *Income class 6 (Low)* | **0.47** | 0.22, 0.72 | 0.57 | 0.31, 0.82 | **0.35** | 0.11, 0.59 | **0.48** | 0.22, 0.73 |
| **Region** | 0.11 | 0.05, 0.22 | 0.02 | 0.01, 0.04 | **0.05** | 0.03, 0.11 | **0.11** | 0.05, 0.22 |

| **SI Table 7 Multivariable logistic regression of improvements in school star ranking between baseline and endline (with economic status of school locations (n = 20, 779)** | | | | | | | | |
| --- | --- | --- | --- | --- | --- | --- | --- | --- |
|  | **Overall School WASH Star Ranking** | | **Water** | | **Sanitation** | | **Hygiene** | |
|  | OR | 95% CI  (Lower, Upper) | OR | 95% CI  (Lower, Upper) | OR | 95% CI  (Lower, Upper) | OR | 95% CI  (Lower, Upper) |
| **MOOC Participation** | | | | | | | | |
| *No school participated* | Ref | | Ref | | Ref | | Ref | |
| *At least 1 teacher enrolled, but no teacher completed* | **1.38** | 1.17, 1.62 | 1.09 | 0.94, 1.28 | 1.11 | 0.95, 1.31 | **1.47** | 1.25, 1.73 |
| *At least 1 teacher completed the MOOC* | **1.46** | 1.31, 1.64 | 1.08 | 0.98, 1.20 | 1.09 | 0.99, 1.22 | **1.43** | 1.28, 1.60 |
| **Type of School** |  |  |  |  |  |  |  |  |
| *Elementary* | Ref | | Ref | | Ref | | Ref | |
| *Secondary* | 1.00 | 0.91, 1.10 | **1.20** | 1.09, 1.31 | **1.13** | 1.03, 1.25 | 0.80 | 0.73, 0.89 |
| **Location of Schools** | | | | | | | | |
| *Urban* | Ref | | Ref | | Ref | | Ref | |
| *Sei-urban* | **1.20** | 1.05, 1.39 | 1.11 | 0.98, 1.26 | 0.92 | 0.81, 1.04 | **1.26** | 1.10, 1.44 |
| *Rural* | 0.82 | 0.64, 1.05 | 0.90 | 0.71, 1.15 | **0.69** | 0.55, 0.88 | 0.89 | 0.70, 1.14 |
| **School Size** | | | | | | | | |
| *Small* | Ref | | Ref | | Ref | | Ref | |
| *Medium* | **1.69** | 1.54, 1.86 | 1.04 | 0.95, 1.14 | **1.55** | 1.41, 1.70 | **1.53** | 1.39, 1.69 |
| *Large* | **1.68** | 1.43, 1.98 | 1.04 | 0.89, 1.21 | **1.42** | 1.21, 1.65 | **1.59** | 1.36, 1.87 |
| *Very Large* | **1.96** | 1.69, 2.28 | 1.08 | 0.94, 1.24 | **1.36** | 1.18, 1.57 | **1.71** | 1.48, 1.98 |
| **Economic status of school locations** | | | | | | | | |
| *Income class 1 (High)* | Ref | | Ref | | Ref | | Ref | |
| *Income class 2* | 1.07 | 0.98, 1.19 | 1.07 | 0.97, 1.17 | **1.09** | 1.00, 1.20 | 1.06 | 0.96, 1.17 |
| *Income class 3* | 1.01 | 0.92, 1.09 | 0.92 | 0.85, 0.99 | 0.97 | 0.89, 1.06 | 0.99 | 0.91, 1.09 |
| *Income class 4* | **1.17** | 1.07, 1.28 | **1.14** | 1.05, 1.25 | 1.05 | 0.96, 1.14 | **1.14** | 1.05, 1.25 |
| *Income class 5* | 0.96 | 0.85, 1.09 | **1.17** | 1.06, 1.31 | 0.96 | 0.86, 1.07 | 0.97 | 0.86, 1.09 |
| *Income class 6 (Low)* | **2.08** | 1.21, 3.59 | **2.09** | 1.24, 3.53 | 1.51 | 0.88, 2.58 | **2.01** | 1.18, 3.42 |
| **Region** | 0.56 | 0.28, 1.13 | 0.06 | 0.02, 0.14 | 0.22 | 0.51, 0.92 | **0.49** | 0.24, 0.98 |

| **SI Table 8 Multivariable logistic regression of improvements in school star ranking from zero between baseline and endline (with economic status of school locations** | | | | | | | | |
| --- | --- | --- | --- | --- | --- | --- | --- | --- |
|  | **Overall School WASH Star Ranking**  (n=19, 018) | | **Water**  (n=3, 219) | | **Sanitation**  (n=11, 771) | | **Hygiene**  (n= 18, 751) | |
|  | OR | 95% CI  (Lower, Upper) | OR | 95% CI  (Lower, Upper) | OR | 95% CI  (Lower, Upper) | OR | 95% CI  (Lower, Upper) |
| **MOOC Participation** | | | | | | | | |
| *No school participated* | Ref | | Ref | | Ref | | Ref | |
| *At least 1 teacher enrolled, but no teacher completed* | **1.45** | 1.22, 1.73 | 1.38 | 0.69, 2.72 | **1.51** | 1.14, 1.99 | **1.49** | 1.25, 1.78 |
| *At least 1 teacher completed the MOOC* | **1.91** | 1.68, 2.17 | 1.26 | 0.81, 1.99 | **1.54** | 1.27, 1.85 | **1.79** | 1.57, 2.03 |
| **Type of School** |  |  |  |  |  |  |  |  |
| *Elementary* | Ref | | Ref | | Ref | | Ref | |
| *Secondary* | 0.94 | 0.84, 1.03 | **1.28** | 0.89, 1.99 | **2.90** | 2.37, 3.54 | **0.75** | 0.68, 0.83 |
| **Location of Schools** | | | | | | | | |
| *Urban* | Ref | | Ref | | Ref | | Ref | |
| *Sei-urban* | **1.25** | 1.08, 1.44 | **1.63** | 1.15, 2.23 | 0.94 | 0.78, 1.11 | **1.34** | 1.17, 1.55 |
| *Rural* | 0.82 | 0.63, 1.07 | 1 | -- | 0.82 | 0.56, 1.18 | 0.93 | 0.71, 1.20 |
| **School Size** | | | | | | | | |
| *Small* | Ref | | Ref | | Ref | | Ref | |
| *Medium* | **1.82** | 1.64, 2.01 | **1.70** | 1.11, 2.61 | **2.01** | 1.73, 2.33 | **1.58** | 1.42, 1.74 |
| *Large* | **1.86** | 1.56, 2.22 | 1.28 | 0.58, 2.78 | **2.87** | 2.08, 3.96 | **1.66** | 1.39, 1.97 |
| *Very Large* | **2.19** | 1.87, 2.57 | **8.06** | 1.08, 59.89 | **4.28** | 2.92, 6.27 | **1.86** | 1.59, 2.18 |
| **Economic status of school locations** | | | | | | | | |
| *Income class 1 (High)* | Ref | | Ref | | Ref | | Ref | |
| *Income class 2* | 1.09 | 0.98, 1.22 | 1.31 | 0.96, 1.78 | **1.29** | 1.14, 1.48 | 1.10 | 0.99, 1.22 |
| *Income class 3* | 1.08 | 0.99, 1.19 | 1.30 | 0.97, 1.73 | **1.15** | 1.03, 1.29 | 1.06 | 0.97, 1.16 |
| *Income class 4* | **1.22** | 1.12, 1.35 | 1.07 | 0.81, 1.41 | **1.22** | 1.08, 1.38 | **1.21** | 1.10, 1.34 |
| *Income class 5* | 0.99 | 0.87, 1.13 | 1.38 | 0.95, 2.01 | 0.99 | 0.85, 1.15 | 1.02 | 0.90, 1.16 |
| *Income class 6 (Low)* | **2.32** | 1.31, 4.09 | 4.44 | 0.58, 2.31 | 1.39 | 0.66, 2.88 | **2.15** | 1.24, 3.76 |
| **Region** | 0.74 | 0.03, 1.51 | 0.25 | 0.08, 0.75 | 0.56 | 0.27, 1.14 | 0.59 | 0.29, 1.21 |
